# Evaluating Text-to-Image Generated Photorealistic Images of Human Anatomy

**DOI:** 10.1101/2024.08.21.24312353

**Authors:** Paula Muhr, Yating Pan, Charlotte Tumescheit, Ann-Kathrin Kübler, Hatice Kübra Parmaksiz, Cheng Chen, Pablo Sebastián Bolaños Orozco, Soeren S. Lienkamp, Janna Hastings

## Abstract

**Background:** Generative AI models that can produce photorealistic images from text descriptions have many applications in medicine, including medical education and synthetic data. However, it can be challenging to evaluate and compare their range of heterogeneous outputs, and thus there is a need for a systematic approach enabling image and model comparisons.

**Methods:** We develop an error classification system for annotating errors in AI-generated photorealistic images of humans and apply our method to a corpus of 240 images generated with three different models (DALL-E 3, Stable Diffusion XL and Stable Cascade) using 10 prompts with 8 images per prompt. The error classification system identifies five different error types with three different severities across five anatomical regions and specifies an associated quantitative scoring method based on aggregated proportions of errors per expected count of anatomical components for the generated image. We assess inter-rater agreement by double-annotating 25% of the images and calculating Krippendorf’s alpha and compare results across the three models and ten prompts quantitatively using a cumulative score per image.

**Findings:** The error classification system, accompanying training manual, generated image collection, annotations, and all associated scripts are available from our GitHub repository at https://github.com/hastingslab-org/ai-human-images. Inter-rater agreement was relatively poor, reflecting the subjectivity of the error classification task. Model comparisons revealed DALL-E 3 performed consistently better than Stable Diffusion, however, the latter generated images reflecting more diversity in personal attributes. Images with groups of people were more challenging for all the models than individuals or pairs; some prompts were challenging for all models.

**Interpretation:** Our method enables systematic comparison of AI-generated photorealistic images of humans; our results can serve to catalyse improvements in these models for medical applications.

**Funding:** This study received support from the University of Zurich’s Digital Society Initiative, and the Swiss National Science Foundation under grant agreement 209510.

**Research in context:** *Evidence before this study:* The authors searched PubMed and Google Scholar to find publications evaluating text-to-image model outputs for medical applications between 2014 (when generative adversarial networks first become available) and 2024. While the bulk of evaluations focused on task-specific networks generating single types of medical image, a few evaluations emerged exploring the novel general-purpose text-to-image diffusion models more broadly for applications in medical education and synthetic data generation. However, no previous work attempts to develop a systematic approach to evaluate these models’ representations of human anatomy.

*Added value of this study:* We present an anatomical error classification system, the first systematic approach to evaluate AI-generated images of humans that enables model and prompt comparisons. We apply our method to a corpus of generated images to compare state of the art large-scale models DALL-E 3 and two models from the Stable Diffusion family.

*Implications of all the available evidence:* While our approach enables systematic comparisons, it remains limited by subjectivity and is labour-intensive for images with many represented figures. Future research should explore automation of some aspects of the evaluation through coupled segmentation and classification models.

## 1. Introduction

Generative AI is poised to transform medicine through a range of potential future applications^1–3^. However, the evaluation of AI-generated outputs poses significant challenges^4^, as generated content is heterogeneous and subject to biases^5,6^. While the bulk of research into the impact of generative models in medicine has thus far focused on the generation of natural language texts, multi-modal models such as those that generate images promise an even greater impact^3^. Here, we focus on text-to-image generative models, such as DALL-E and Stable Diffusion, that enable the creation of photorealistic images from natural language descriptions. Applications of these models in medicine include generating representative illustrations for medical education, and creating synthetic data for a variety of scientific purposes in contexts where real-world images are insufficient, lacking, or unavailable due to privacy concerns. While task-specific models using Generative Adversarial Networks (GANs)^7,8^ have been available for some time, diffusion-based text-to-image models are distinguished by their generality and ease of use^9^.

The potential of these text-to-image models in medicine is already being evaluated, for example to generate accurate anatomical illustrations of the human skull, heart and brain^10^, human faces and other body parts with various pathologies^11–13^, and illustrations of resuscitation techniques^14,15^. Their applicability has also been tested for surgical planning and patient consultation in dental surgery^16^ and aesthetic surgery^17^. Moreover, DALL-E 2 has been used to synthesise realistic images for training predictive algorithms in dermatology^18^ and for aphasia assessment^19^. The technology has also been implemented to generate synthetic radiology images, e.g. chest x-rays^20,21^.

It has been observed that a variety of visual errors are common in generated images, in particular, when rendering human hands^22^. Such inadvertent errors may potentially blur the diagnostic boundaries between the anatomical representations of healthy and diseased anatomy, which are essential for downstream applications in medicine. However, thus far, there has been no systematic examination of such errors or their direct comparison across different generative models. To address this gap, our study introduces a novel method of descriptive evaluation of anatomical errors in generated photorealistic images of the human body. Importantly, our focus on assessing model-inherent errors in generative photorealistic imagery of human bodies has no intention of discriminating against people with non-normative bodies. Our assumption is that understanding the errors that these models make on the kinds of everyday images they were trained on will be essential for future evaluation of the errors these models make when generating domain-specific medical images.

## 2. Methods

### 2.1. Development of the Anatomical Error Classification System

We developed the anatomical error classification system iteratively, as extensively detailed in Supplementary Method 1. In brief, we generated and annotated a collection of photorealistic images sampled from three models and multiple prompts, then applied our initial classification system to compute inter-rater reliability, as well as to compare models and prompts, followed by improving the system iteratively. The first iteration of the classification system defined five types of errors (missing, extra, configuration, orientation and proportion), and five anatomical regions (torso, limbs, feet, hands and face). In the second iteration, we introduced a proportion-based quantification of the errors in each image (Equation 1 below). In the final iteration, our quantitative system was extended with a weighting of errors by severity across three error severity levels (‘a’-low, ‘b’-medium and ‘c’-severe) along with a cumulative error severity score per image (Equation 2 below).

### 2.2. Scores for Quantitative Comparisons

#### Equation 1 (Proportion of Errors)

For each picture, each body part, each error type and each error severity, we define the proportion of errors as the number of body parts *i* that present with the error type *j* with the corresponding error severity 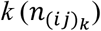 divided by the number of body parts present, or the number of body parts that should be present given the number of people and what is or should be visible in the picture (*m*_*ij*_):

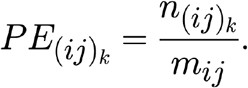

#### Equation 2 (Cumulative Score)

Let 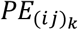 be the number of body parts *i* that exhibit an error of type *j* of error severity *k*. Let *w*_*a*_, *w*_*b*_, *w*_*c*_ be the error severity weights that correspond to the error severity a, b, c, respectively (we used 0.2 for ‘a’, 0.5 for ‘b’, and 1 for ‘c’, to down-weight less severe errors). Then, the cumulative score *C*_*x*_ of image *x* is defined as:

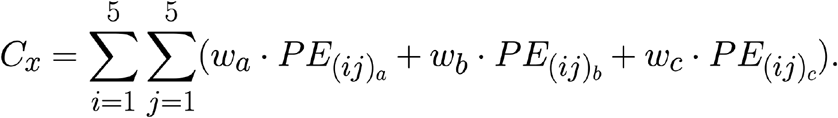

### 2.3. Generating the Image Dataset

We used three models: DALL-E 3 (a commercial model), Stable Diffusion XL and Stable Cascade (two models from the non-commercial Stable Diffusion family). We designed 10 prompts, covering a variety of visual scenarios presenting either individuals, couples or groups of about five people in dynamic action and mutual interaction across heterogeneous everyday settings. The ten prompts were:

1. athlete performing salto
2. person jogging
3. mother (or father) holding baby
4. couple hugging
5. two men (or women) wrestling in an arena
6. old couple in sauna
7. physician examining patient
8. people eating pizza
9. five people sunbathing on a beach
10. five people playing volleyball

For each prompt, we generated 8 images. We focused on obtaining a gender-balanced dataset: when a generative model showed implicit bias toward a particular gender, we explicitly prompted to counter this bias. We also focused on obtaining a dataset that covered a range of age groups, from babies to older individuals, and included diverse ethnicities, although most images nevertheless represent Caucasian individuals.

### 2.4. Allocating Image Annotations

Our annotation team consisted of four annotators. To be able to assess the inter-rater agreement between the annotators, we submitted 25% of our dataset (60 images) to double annotation. This resulted in 300 annotations that were equally distributed across the annotators. To ensure that the three generative models, the 10 prompts, and the 60 double-annotated images were randomly distributed across the 4 annotators, we automated the distribution (script available on our GitHub repository^23^).

### 2.5. Assessing Inter-Rater Reliability

We assessed the inter-rater agreement by calculating the Krippendorff’s alpha coefficient^24^ using the Python package fast-krippendorff. We applied Krippendorff’s alpha to three variations of our annotation results: 1) the cumulative error score for each image; 2) an overall error severity for each image (low, medium, high); and 3) a vector of 25 entries for each category and body part with 0 for no error and 1 for an error. We determined an overall error severity by using the cumulative score to categorise each image based on quantiles of the error distribution of each individual annotator: with the 0.5 quantile as a threshold for overall error level *low*, 0.75 for overall error level *medium*, above 0.75 as overall error level *high*.

### 2.6. Quantitative Model Comparison

We applied the error classification system to several different comparisons using our image dataset: between models, between error types, between anatomical regions, and between individual vs. group prompts. For these comparisons, we used the cumulative score and the aggregate counts per error severity to compare the models.

## 3. Results

### 3.1. The Anatomical Error Classification System

The anatomical error classification system supports the identification and quantification of different anatomical errors in photorealistic images of humans based on visual analysis of individual images.

The system (Figure 1) consists of three components: (1) five anatomical regions – face, torso, hands, limbs and feet (Figure 1A); (2) five error types – proportion, extra, orientation, configuration, and missing (Figure 1B); and (3) a quantification of error severity and method of aggregation. The error types are:

**Figure 1:**
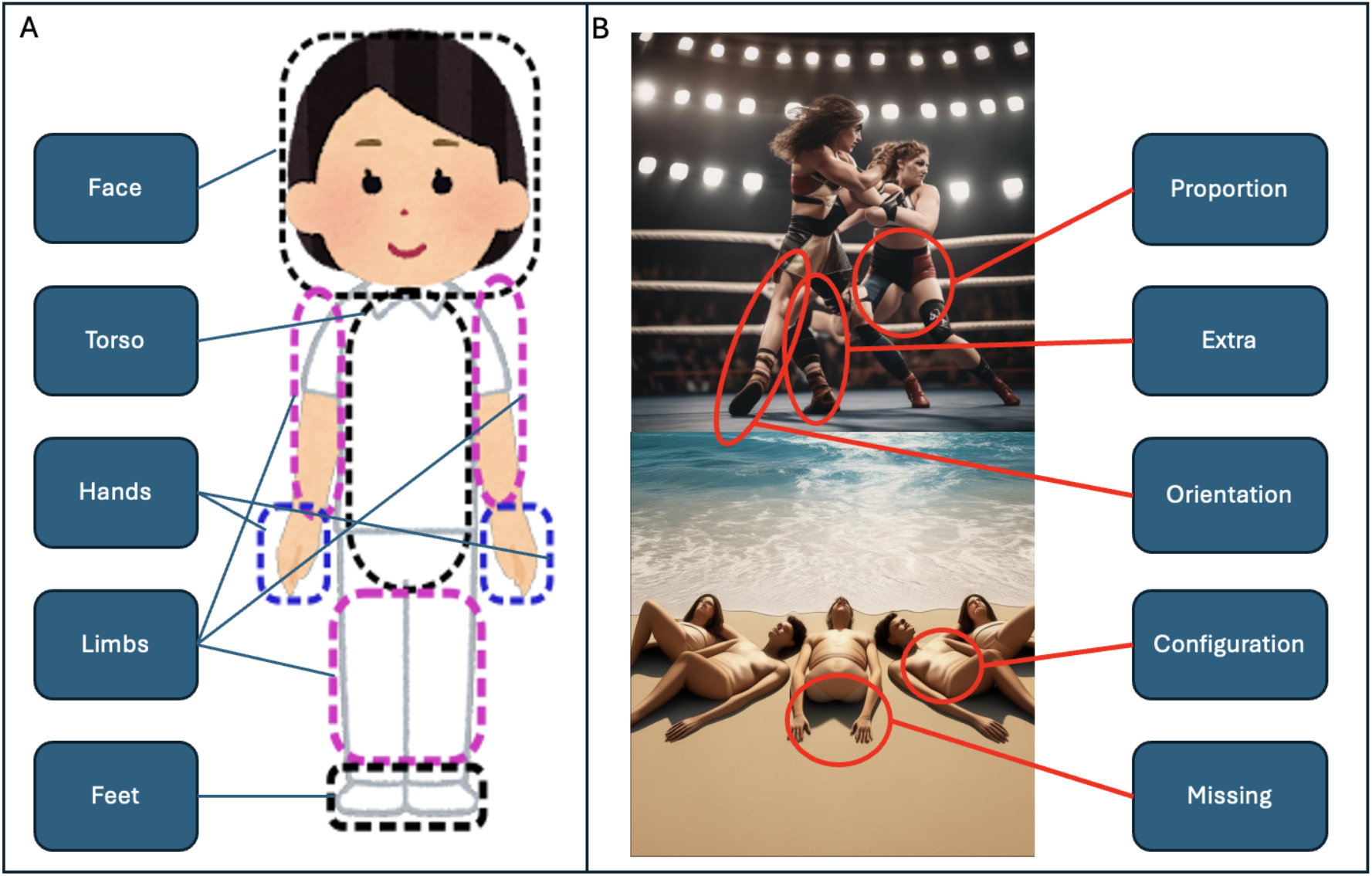
The Anatomical Error Classification System – illustration of visual components (A) – anatomical regions; (B) – types of error.

1. *Proportion errors:* misshaped or disproportionate body parts (e.g., limbs that are too short for the body).
2. *Extra errors:* additional body parts (e.g., a hand with six fingers)
3. *Orientation errors:* anatomically implausible orientations of body parts (e.g., the upper and lower body facing in opposite directions),
4. *Configuration errors:* body parts that are disjointed (e.g. a hand disconnected from the body), displaced (e.g., a hand connected to the chest), or fused in ways that make their differentiation challenging (e.g., a hand merged with a fork)
5. *Missing errors:* absent body parts (e.g., an arm without a hand)

We developed an annotation manual (see our repository^23^) to mitigate inconsistent decisions among annotators. The manual introduces and explains the system using annotated examples. It instructs the annotators to only consider the main anatomical errors in generated images evident at first glance, while disregarding minor errors that require zooming in to detect.

### 3.2. The Image Dataset and Qualitative Evaluation

Our three models, 10 prompts and 8 images per prompt resulted in a dataset of 240 generated images, available from our repository^23^. Example images are shown in Figure 2.

**Figure 2:**
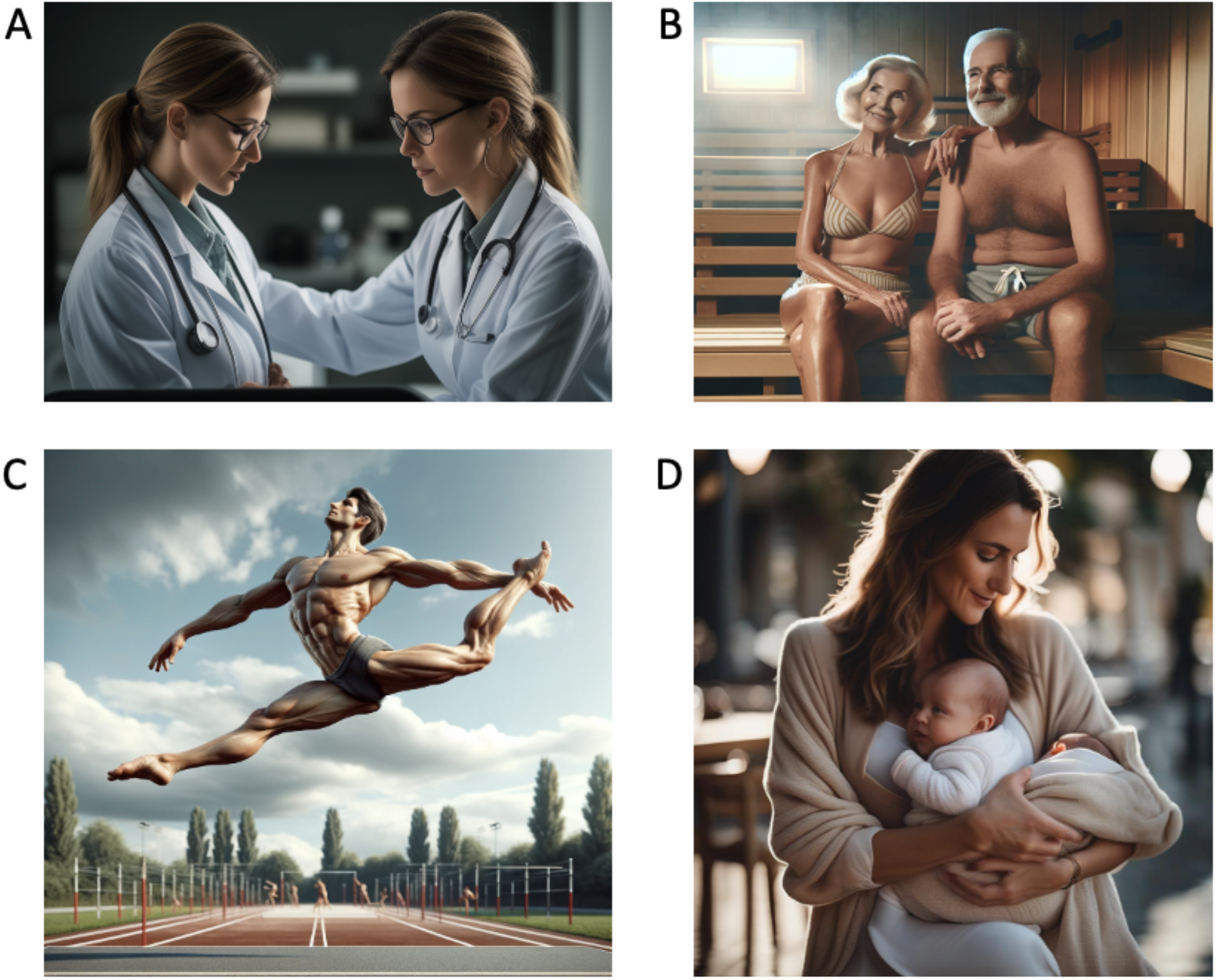
Selected example images from the generated image dataset. (A) physician examining patient; (B) old couple in sauna; (C) athlete performing salto; (D) mother holding baby.

While developing our image dataset, we noticed that some of the designed prompts proved to be challenging for some of the models. For example, the prompt about an aged couple in a sauna repeatedly led to an error in DALL-E 3 and even to temporary blocking of the account for a few hours without any explanation. We had a similar experience with the prompt that instructed DALL-E 3 to generate two women wrestling in an arena, but not when prompting for wrestling men. The other two models did not have any problems with these prompts. Initially, we planned to use the prompt ‘a couple kissing.’ However, DALL-E 3 declared this forbidden content, so we adjusted the prompt to ‘a couple hugging.’

We also noticed that when prompting the models to generate images that included two individuals, these individuals often looked almost identical, like twin or digital copies of each other. Another pattern that became apparent was that doctors always had a stethoscope, a detail that was impossible to avoid even when using negative prompting. Additional stereotypes included mothers being predominantly young and always having long hair, whereas older people, despite having somewhat wrinkled faces, were generated with bodies having impeccably smooth skin. Relatedly, across all models, the prompt ‘a couple hugging’ resulted in images of heteronormative couples.

Ethnic diversity was considerably more present in Stable Diffusion XL and Stable Cascade than DALL-E 3. When prompted to generate an image of a person without any explicit specification of gender, Stable Diffusion XL and Stable Cascade generated more gender-diverse images, whereas we had to prompt DALL-E 3 explicitly about gender aspects to generate a gender-balanced dataset.

### 3.3. Annotation Results and Quantitative Evaluation

The distribution of cumulative scores for all images and all annotators is shown in Figure 3A, and the distribution of overall per-annotator error severity levels by model is shown in Figure 3B.

**Figure 3:**
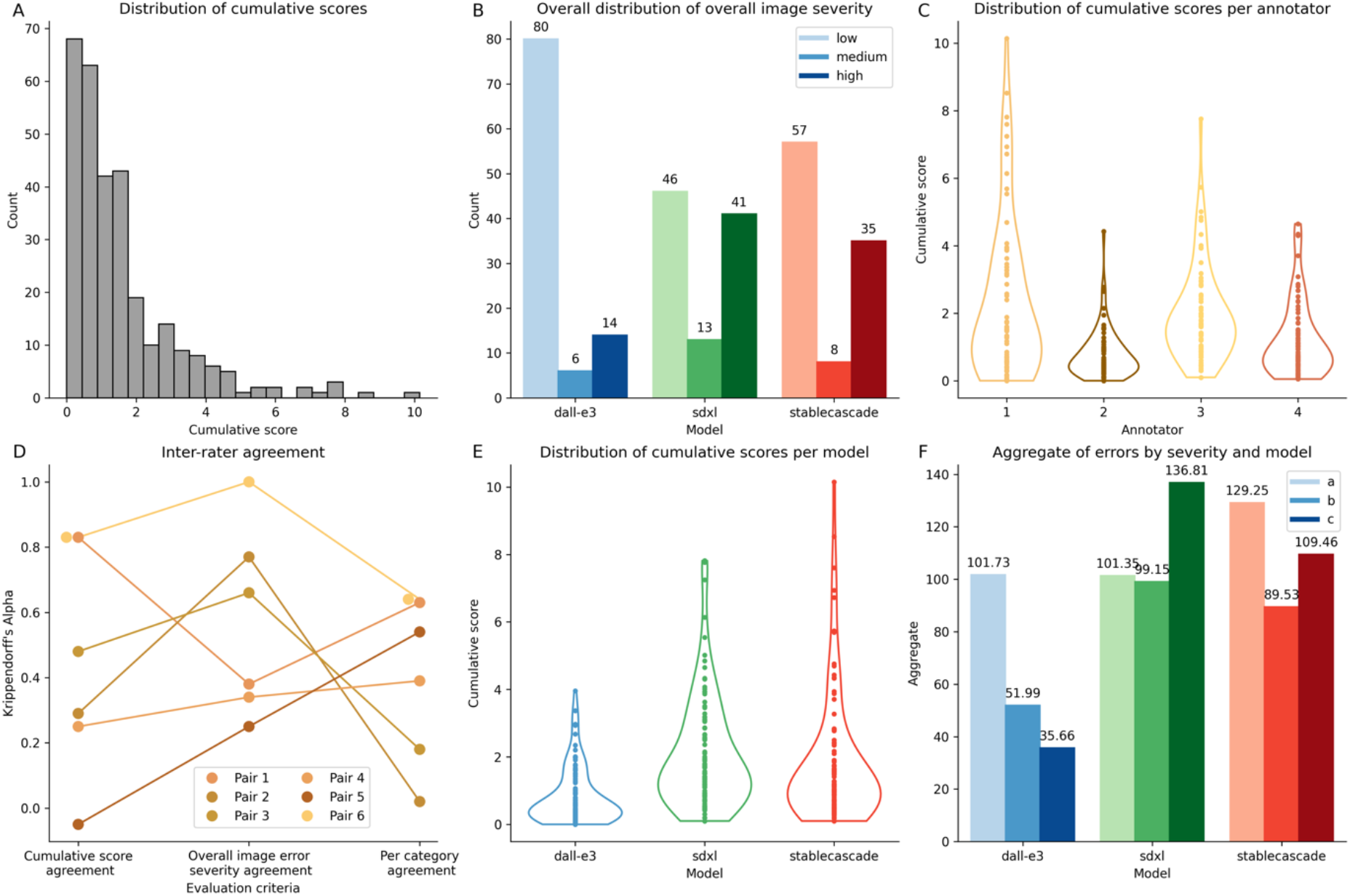
A – Overall cumulative score distributions for annotation dataset. B – Distributions of overall image severity per model. C – Distribution of cumulative scores per annotator. D – pair-wise inter-rater agreement for the pairs of annotators across metrics. E – Distribution of cumulative scores per model. F – Aggregate error scores by severity per model.

The distribution of scores for each of the four annotators (Figure 3C) reveals potential inter-rater differences in annotation styles which reflect the subjectivity of the task. Figure 3D shows the inter-rater agreement for individual annotator pairs for the three different metrics: cumulative score agreement, overall per-annotator error severity agreement, and agreement per category. The average Krippendorff’s alpha coefficient for the cumulative score agreement, the overall error severity agreement and the binary category agreement are 0.44, 0.57 and 0.45, respectively. These are above random, but reflect a relatively poor level of agreement, which may be expected given the complexity and subjectivity of the rating task.

Figure 3E shows the distribution of cumulative scores for images across models. DALL-E 3 has the lowest cumulative error scores overall (mean 0.82, variance 0.73; Welch t-test statistic -6.72 and - 4.67; p-value 3.72E-10 and 7.12E-6; comparing DALL-E 3 to SDXL and Stable Cascade, respectively). Stable Cascade and SDXL were not significantly different (Welch t-test statistic of -1.05 and p-value 0.3), although we observed a slightly lower mean for SDXL but higher variance (mean of 2.1 and 1.8, variance of 2.72 and 3.69 for Stable Cascade and SDXL, respectively).

Next, we compared the aggregate counts per error severity scores (‘a’, ‘b’, and ‘c’) across models (Figure 3F). While DALL-E 3 has a similar number of errors with severity ‘a’ as SDXL (101.73 and 101.35, respectively), it has much fewer errors of severity ‘b’ or ‘c’ than the other models (severity b: 51.99, 99.15, 89.53 for DALL-E 3, Stable Cascade, SDXL, respectively, severity c: 35.66, 136.81, 109.46 for DALL-E 3, Stable Cascade, SDXL, respectively). Furthermore, SDXL has the highest number of severe ‘c’ errors.

Next, we compared cumulative scores and aggregate error counts per error severity for each prompt (Figure 4). Certain prompts were more challenging and error-prone than others. For example, the prompt “five people sunbathing on a beach”, led to notably higher cumulative error score (mean 3.96, variance 6.1) than the prompt “mother or father holding a baby” (mean 0.82, variance 0.42) (Figure 4A). The difference is statistically significant (Welch t-test statistic 6.74; p-value 1.12E-7). The former prompt not only had the highest cumulative error score but also a significant number of ‘c’ (severe) errors (Figure 4B). DALL-E 3, SDXL, and Stable Cascade exhibited numerous ‘a’, ‘b’, and ‘c’ errors, with ‘c’ errors (179.72) exceeding the cumulative counts of ‘a’ (71.55) and ‘b’ (86.67) errors.

**Figure 4:**
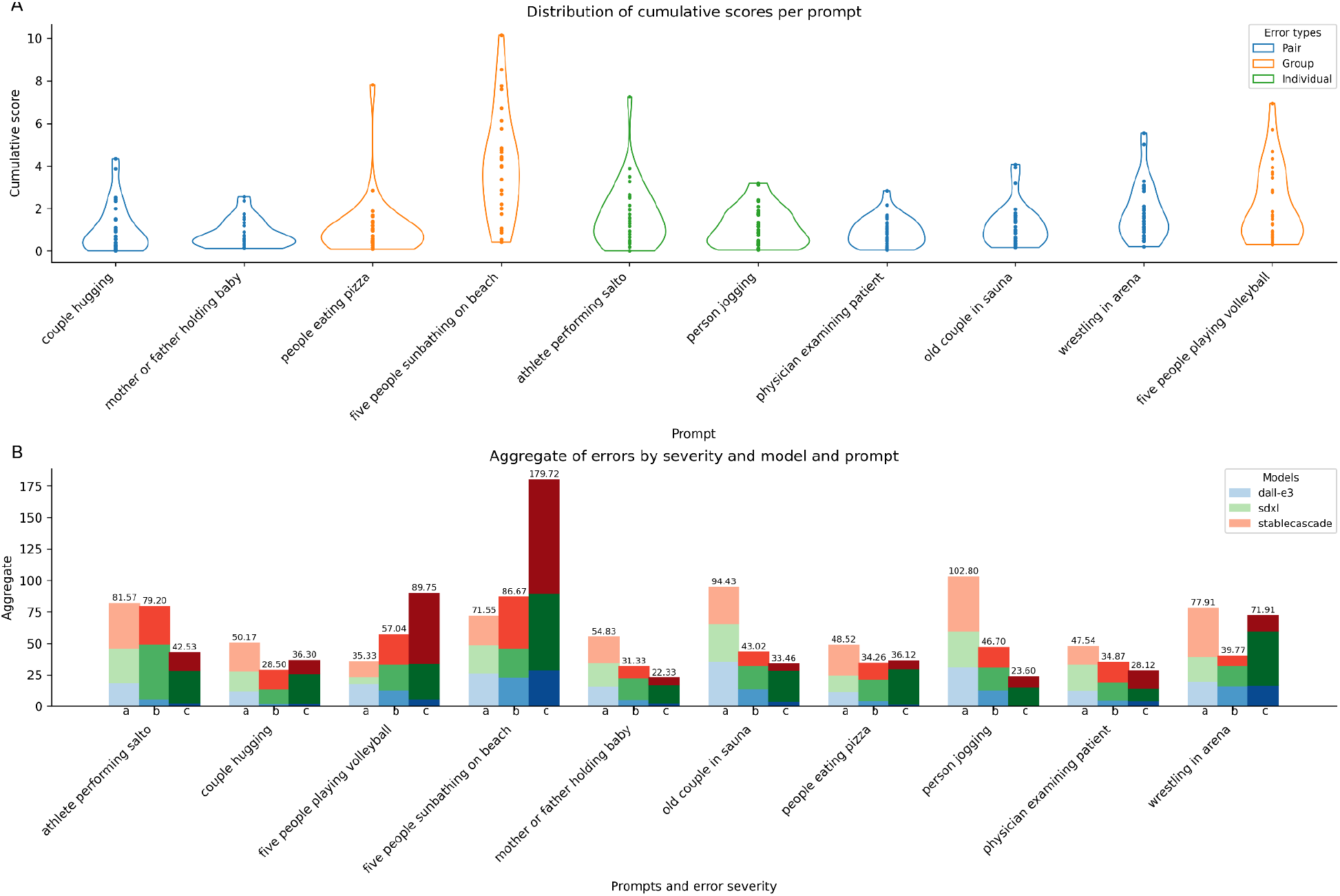
Errors per prompts. A - Cumulative score. B - Error aggregates per severity

In general, prompts involving groups of people, such as “five people sunbathing on the beach” and “five people playing volleyball,” not only had higher cumulative error scores but also exhibited a more dispersed score distribution (Figure 5A). In terms of the per-severity error counts (Figure 5B), prompts featuring groups of people had more errors overall, with a higher proportion of ‘c’ errors compared to prompts involving one or two people, although “people eating pizza” was an exception. Across all prompts, DALL-E 3 consistently demonstrated the best performance, with lower overall error counts and fewer severe ‘c’ errors. Aside from “five people sunbathing on the beach,” DALL-E 3 showed a pattern where ‘a’ error counts exceeded those of ‘b’ and ‘c’ errors. Stable Diffusion XL and Stable Cascade performed similarly, however, aside from “five people sunbathing on the beach” and “five people playing volleyball,” Stable Diffusion Cascade had lower total error counts than Stable Diffusion XL and generally fewer ‘c’ errors.

**Figure 5:**
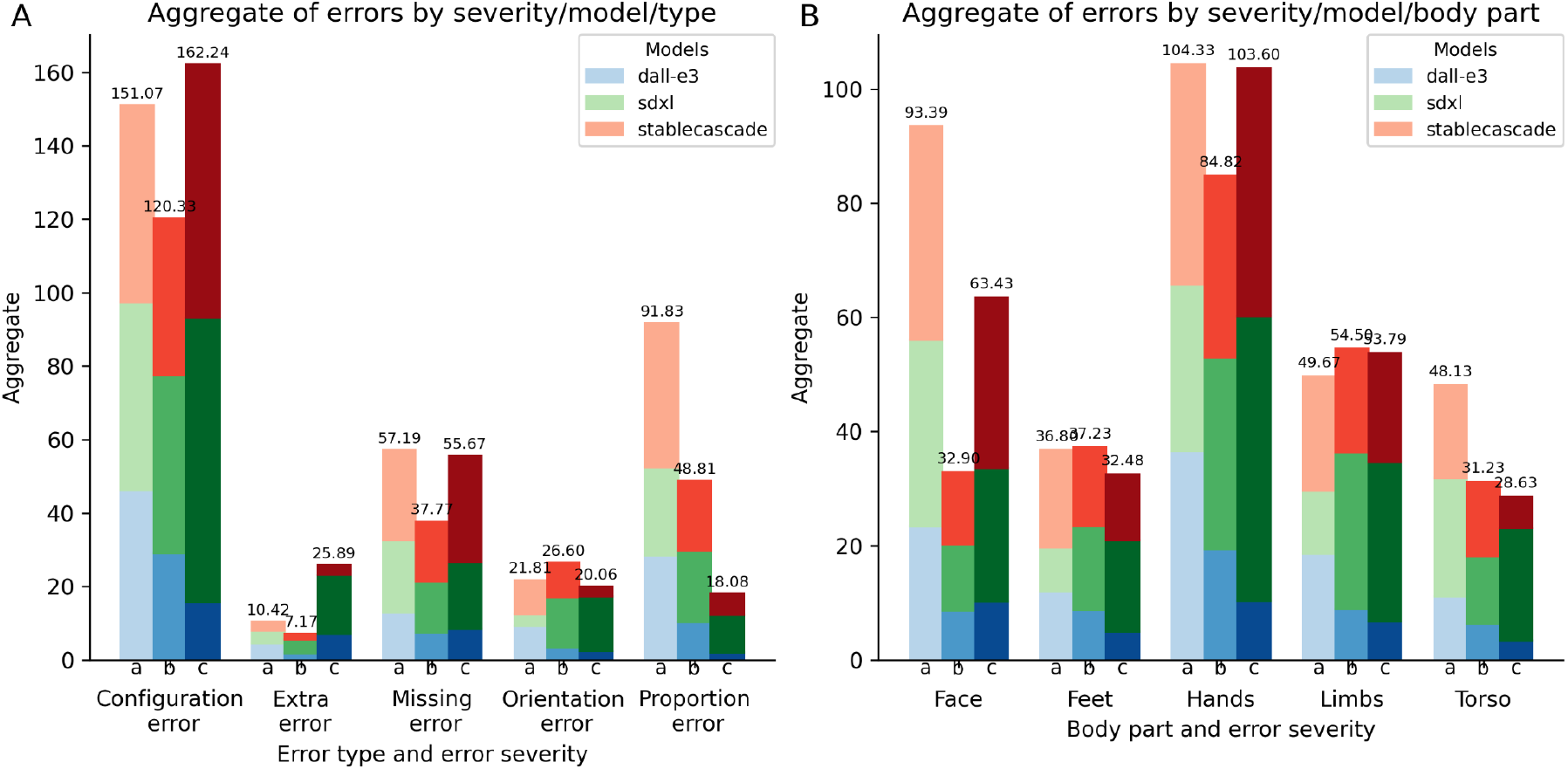
Aggregates of errors by error type (A) and body part (B)

For all models, configuration errors are the most prominent, with the count of ‘a’, ‘b’, and ‘c’ levels far exceeding those of the other error types (Figure 5A). Moreover, the number of ‘c’ errors surpasses both ‘a’ and ‘b’ errors, indicating that not only are configuration errors more common, but also often quite severe. Missing errors follow, with similar counts of ‘a’ and ‘c’ errors. Surprisingly, extra errors rank last in the total count. Nonetheless, these errors are more severe when they occur. Conversely, proportion errors, despite their higher overall count, are predominantly ‘a’ errors. For instance, many images generated by DALL-E 3 feature exaggerated muscle or body proportions without missing or extra body structures. Regarding body parts, hands are the most problematic anatomical region (Figure 5B). Face and limbs follow, with feet and torso being the least problematic.

## 4. Discussion

Generative text-to-image models have the potential for wide-ranging future applications in the medical domain, including for privacy-preserving synthetic data generation. However, it is necessary to better understand the limitations of such models and qualitatively and quantitatively assess the errors they make in generating images of human anatomy. Assessing generated content is challenging and subjective, and to the best of our knowledge this is the first attempt to create a systematic approach with broad applicability.

There are several limitations to our method. As tested through double annotations and quantifications of inter-rater agreement in our sample, there is a subjective aspect in how the errors are identified in the images, particularly regarding their varying severity levels. The introduction of a manual aims to reduce subjectivity by defining operative rules for how to apply the method. We also recommend training and discussions to mutually align annotators’ potentially subjective interpretations of error severities. However, even with the manual and training, it is impossible to fully exclude subjectivity from each annotator’s decisions on what counts as an anatomical error in generated images and error severity. This is because such decisions are necessarily influenced by multiple subjective factors, such as the annotator’s implicit assumptions about what counts as ‘normal’ anatomy or which body parts should be visible in an image from a particular perspective.

There are two other limitations to our method. First, for images that show larger groups of multiple individuals, the application of our method’s manual error counting becomes impractical since the number of anatomical parts that must be visually examined for errors becomes too large and the body parts too small to be assessed without zooming in. For this reason, in our image dataset, we prompted for a maximum of five figures (resulting in images containing up to seven figures given the variability in outputs). Second, for practical reasons, we limited the error types to five, as increasing the number of types would have led to a significant increase in error categories (number of error types x number of anatomical regions), thus potentially overwhelming the annotators. This, however, meant that some error types (e.g. configuration and proportion errors) are more broadly defined than others (e.g. missing, extra, and orientation errors).

The qualitative findings that we observed during the process of generating our dataset with the ten prompts are fully aligned with and further contribute to the ongoing discussion in the current literature around the implicit gender and ethnic biases of generative models ^25–28^. By prompting for older people in a sauna, we have also demonstrated an implicit ageism bias in these models, as images of older individuals were often rendered with unrealistically youthful bodies, a type of bias that has to our knowledge not previously been reported in the literature.

Perhaps unsurprisingly, in our quantitative comparison the commercial model DALL-E 3 performed better than the competing non-commercial alternatives. However, this result has to be balanced alongside qualitative observations of problems that were not covered by our anatomical error classification system, such as the ethnic bias, as well as the observed “twinning” (multiple people having the same facial features) and “hyper-idealisation” (overly perfect and unrealistic representations) of human figures, which was more prominent in DALL-E.

More surprising was the result of the diachronic comparison of two versions of Stability AI’s models, the more recent Stable Cascade and the earlier Stable Diffusion XL. In our dataset, the quantitative comparison of errors showed that Stable Cascade generated a smaller amount of serious errors than Stable Diffusion XL but a comparable amount of moderate and even a larger amount of minor errors. We presume that the reason for this relative similarity of performance can be attributed to the fact that these two models use different architectures and that the Stable Cascade was primarily developed to optimise the speed, efficiency and flexibility for fine-tuning of image generation, rather than to substantially reduce errors in the generated images^29^.

Interestingly, in developing a range of prompts that covered different everyday scenarios, we discovered that some prompts challenged all the models. For example, “five people sunbathing on a beach” led to a surprisingly high number of severe errors in all three models. Our quantitative findings also reveal that prompts that describe less common bodily constellations, such as two people wrestling in an arena or a single individual performing a salto, also challenge all the models we tested, resulting in an increase of anatomical errors compared to less challenging prompts, such as a mother or father holding a baby. This is in line with previous results evaluating generative AI models which have shown that such models’ performance drops for prompts corresponding to scenarios less present in the training dataset^30^.

A limitation of our findings is that our dataset was relatively small and only contained images generated by three text-to-image models using ten prompts. In the future, a more comprehensive comparison across further models using a larger number of prompts that cover a more heterogeneous range of everyday scenarios would be desirable. However, such comparison would require a larger team of annotators, which, in turn, could potentially exacerbate the problem of limited inter-rater agreement when applying a method that necessarily entails a subjective element of image evaluation. Thus, we also plan to explore approaches that use additional model types such as segmentation to attempt to partially automate parts of the error detection and classification system.

## 5. Conclusions

Generative AI is already having a significant impact in society and in medicine but developing a better understanding of the strengths and limitations of such models requires new methods to evaluate their outputs. Here, we report a novel approach to evaluate errors in the images of human anatomy from such models and apply our approach to compare models and their outputs. Our approach was developed with potential medical applications in mind, to enable systematically differentiating medically meaningful anatomical pathologies from model-driven unintended visual errors, without discriminating against persons with non-normative anatomies. We also foresee broader potential usage in non-medical contexts as generative AI images of humans become more prominent across wide-ranging applications. Additionally, our method could support academic research to assess and compare implicit visual biases of different generative models, supporting the improvement and optimisation of such models in the future.

## Data Availability

All data provided in the manuscript are available online at https://github.com/hastingslab-org/ai-human-images.

## Author contributions

JH initiated, conceptualised and led the project. PM conceptualised the project, led the literature review and wrote the initial draft of the manuscript. YP, AKK, HKP, PSBO and JH developed the anatomical error classification system. PM generated the final image dataset. CT, AKK, PM and YP annotated the final image dataset. CT, YP and AKK analysed the data and generated the plots. SL clinically advised the development of the anatomical error classification system. All authors contributed to revising and editing the manuscript and all authors agree to the current form.

## Acknowledgements

The project was supported by funding from the University of Zurich’s Digital Society Initiative, and by the Swiss National Science Foundation under grant agreement 209510.

