## Supplementary Material for "Evaluating Text-to-Image Generated Photorealistic Images of Human Anatomy"

### Supplementary Methods and Figures

#### Supplementary Methods

Supplementary Method 1. Developing the Error Classification System

*1.1. Generating a Preliminary Image Collection*

To develop a classification system tailored to evaluating technical errors in the photorealistic generative imagery of human anatomy, we started by selecting three different text-to-image generators: DALL-E 3, Stable Diffusion 1.5 and Stable Diffusion XL. Our choice aimed to include both proprietary (DALL-E) and open-source generators (Stable Diffusion), as well as different versions of one model (Stable Diffusion 1.5 and Stable Diffusion XL) in order to take into account its continued optimisation.

Having selected the image-to-text models, we then turned to generating a collection of images by prompting these models. First, we performed an exploratory image generation in order for the research team to familiarise themselves with the technology and to inform the prompting strategy that was used in the subsequent stages. At this point, each team member generated 5 photorealistic images of humans engaging in everyday situations with each of the three models and brought those images to the group discussions. Based on these discussions, we designed 6 simple prompts. The prompts were specifically chosen to cover different everyday scenarios that included a single individual, a couple, or a group of individuals. This choice allowed us to quantify the presence of mistakes depending on the number of individuals represented in the images. The prompts were also created to cover a variety of scenarios that resulted in visualisations of human bodies in either static (e.g., sitting) or dynamic (e.g., dancing) situations. Moreover, in addition to prompting the models to generate imagery of people in everyday environments (e.g., an apartment), we included other contexts, such as being on a beach. The latter choice allowed us to generate images revealing anatomical structures that typically remain hidden from view in other everyday scenarios. Using our 6 prompts, we generate 10 images per prompt with each of the three models, resulting in an initial collection of 180 images.

*1.2 Two-Stage Development and Testing of the Error Classification System*

Having used the prompts to generate the image material, we then proceeded to develop our error classification system based on the comparative visual analysis of the generated images, which was performed in two stages.

In stage 1, we introduced a preliminary typology of errors to distinguish between different kinds of unintended visual deviations from standard representations of human anatomy. Our goal was to develop categories that are mutually distinct enough so as not to overlap, while at the same time covering the entire range of anatomical errors that we identified in our image sample through visual inspection. Based on the analysis of our image collection, we defined five types of anatomical generation errors:

1. Missing errors, designating an absent body part (e.g., an arm without a hand)
2. Extra errors, referring to the generation of additional body parts (e.g., a hand with six fingers)
3. Configuration errors, encompassing the generation of body parts that are either disjointed (e.g. a hand disconnected from the body), displaced (e.g., a hand connected to the chest instead of the arm), or fused in ways that make the differentiation of individual body parts challenging or impossible (e.g., a hand merged with a fork)
4. Orientation errors, entailing anatomically implausible orientations of various body parts (e.g., the upper and lower body facing in opposite directions), and
5. Proportion errors, comprising distortions arising from misshaped or disproportionate body parts (e.g., limbs that are too short for the visualised body).

In this initial stage, apart from introducing our error typology, we also topographically divided the body into five anatomical regions:

1. Torso,

2. Limbs,
3. Feet (with five toes each),
4. Hands (with five fingers each), and
5. Face.

This topographic division allowed us to evaluate the presence or absence of each of our five error types in each anatomical region separately. The combined use of error types and anatomical subcategories aims to enable the assessment of how well different text-to-image models generate different body parts and potentially identify which types of mistakes more frequently occur in a given anatomical region in a particular text-to-image model.

Next, we turned to evaluating the performance of this preliminary version of our error classification system with a view to optimising it in the subsequent stage of its development. To this end, a team of five annotators applied the system to our initial collection of generative images. To test if our preliminary classification system would achieve sufficient inter-rater agreement, each of the 180 images was annotated by two annotators. We shuffled the images, to ensure that each annotator received images generated by different models and stemming from different prompts. Furthermore, we devised a standardised annotation sheet in Excel. This sheet introduced a temporal sequence in which the annotation is performed both concerning the assessment of different error types and the anatomical regions. According to this predefined sequence, the annotators are required to first look for the missing and extra errors, then proceed with the configuration and orientation errors, and finish with the proportion errors. Relatedly, the annotation sheet imposes the temporal sequence in which, for each error category, the anatomical regions should be scrutinized, starting with the torso and the limbs, then moving on to the hands and the feet, and finally examining the face. In the annotation sheet, the annotator can see the image's identification number, the corresponding prompt, and the model used to generate it. Using the annotation sheet, each annotator marked the presence or absence of a particular error type in a specified anatomical region for each of the images assigned to them.

The comparison of the annotations performed by the different annotators revealed significant disagreements in how to apply our preliminary classification system to the generated images. Based on the analysis of the annotations, we determined that the disagreement primarily stemmed from subjective decisions about the relevance of different error types and their mutual differentiations. We also established that by focusing on the mere presence or absence of a particular error type in a generative image, our preliminary system was unable to differentiate between images that contained a large amount of serious errors (such as those generated by Stable Diffusion 1.5) and those that had a few minor errors (such as those generated by DALL-E 3). Thus, in the next stage, we used these findings to optimise our system.

In stage 2, we introduced a quantification of errors per image. We thus instructed the annotators to first count for each anatomical region separately how often it appears in the image and then evaluate each of the instances of that anatomical region for a particular type of error. For example, if two persons are shown in an image with their entire figures fully visible, then the annotator should separately count each error category for two trunks, four limbs, four hands and four feet, and two faces. The resulting score, expressed as a fraction, denotes how many instances of a particular anatomical region shown in the image exhibit a particular type of mistake (e.g., ¼ limbs with a proportion error). We further introduced into our method the weighting of each error through the assignment of its severity. The grading of the error severity is rated as ‘a’ for a minor error, ‘b’ for a moderate error, and ‘c’ for a severe error. Each severity grade carries a different penalty score: ‘a’ error: 0.2 points; ‘b’ error: 0.5 points; and ‘c’ error: 1 point. At the end of the annotations, the scores for the different error categories are multiplied by the corresponding penalty points depending on the severity grade and then added up to obtain the cumulative error score for the generated image. Thus, through this update, we obtained an evaluation method that allows a more fine-grained quantitative comparison of the quality of images generated by different text-to-image models from the perspective of the models’ limited ability to generate error-free representations of human anatomy.

Finally, we tested our updated classification method for the comparison of images generated by different text-to-image models. We aimed to establish if our classification method would allow us to quantify the level of anatomical errors across the different models based on cumulative scores. Using a proportional score ensures that we can reliably compare images that exhibit different numbers of people.

We performed another annotation round, this time focusing on a small subset of our image collection. Four annotators assessed 25 images with each image being assessed by two annotators. Our statistical analysis of the thus obtained annotation scores revealed that our optimised error classification system was able not only to quantitatively distinguish better performing generative models but also to assess which types of errors and of which severity grade were predominant in each generative model.

**Supplementary Figures**

Figure S1. Aggregate of Errors by Type, Body Part and Prompt

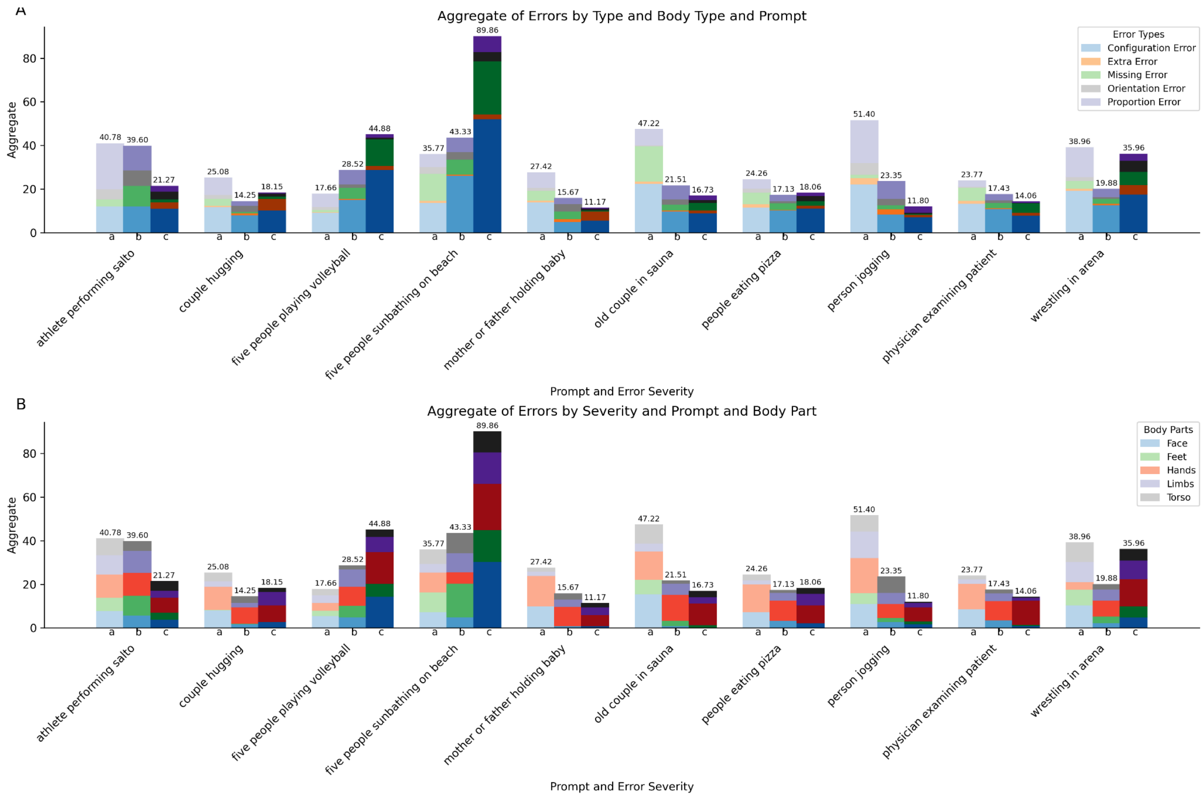


Figure S1 shows the error scores broken down by severity, prompt and body part.

Figure S2. Aggregate of errors by severity, model and prompt for the different error types.

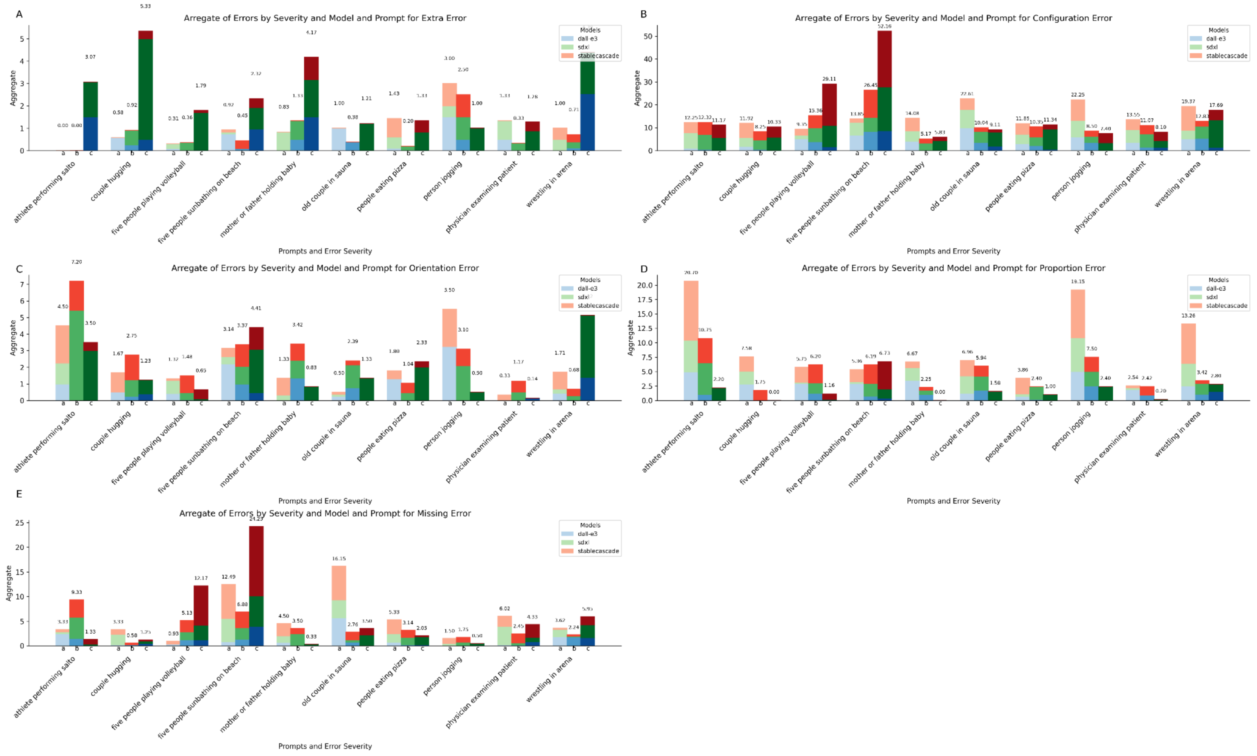

Figure S2 shows the breakdown of errors by severity, model and prompt, for each of the five different error types.

Figure S3. Aggregate of errors by severity and model type per anatomical region.

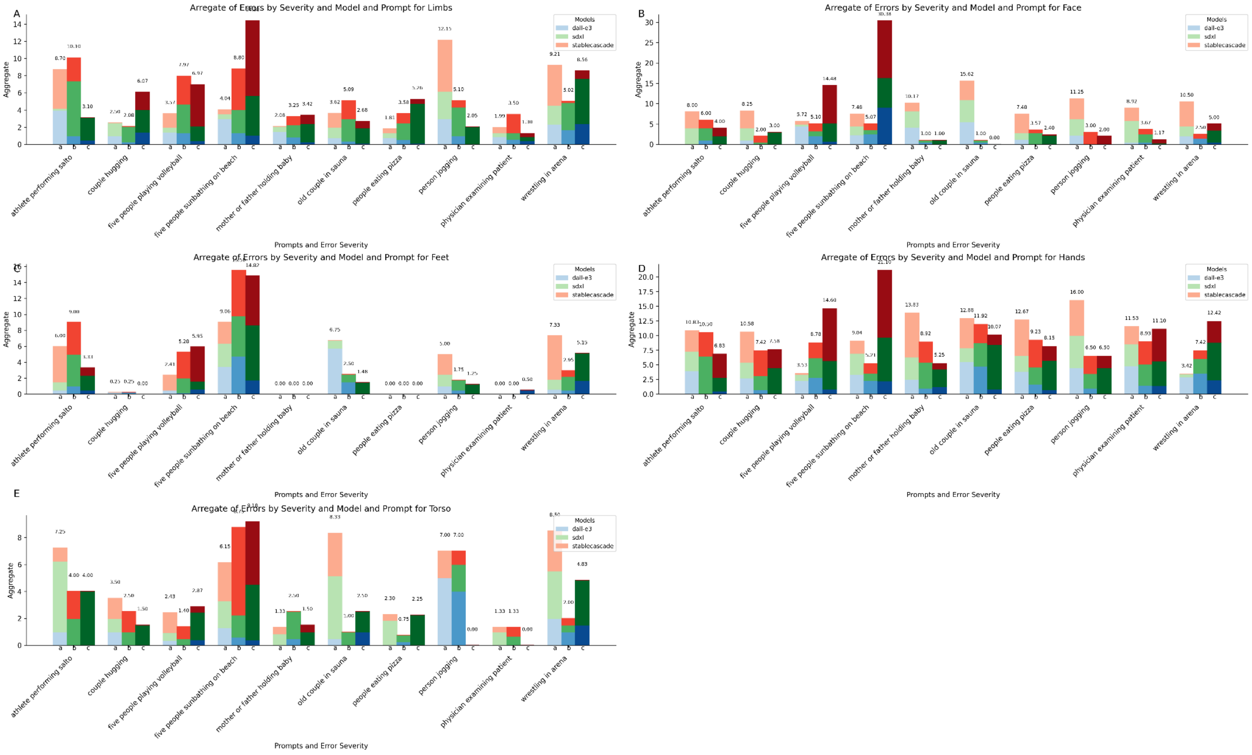


Figure S3 shows the breakdown of errors by severity, model and prompt, for each of the five different anatomical regions.
